# Genetic Contributions to Alcohol Use Disorder Treatment Outcomes: A Genome-wide Pharmacogenomics Study

**DOI:** 10.1101/2021.02.03.21251107

**Authors:** J.M. Biernacka, B.J. Coombes, A. Batzler, J.R. Geske, A.M. Ho, J. Frank, C. Hodgkinson, M. Skime, C. Colby, L. Zillich, S. Pozsonyiova, M-F. Ho, F. Kiefer, M. Rietschel, R. Weinshilboum, S.S. O’Malley, K. Mann, R. Anton, D. Goldman, V.M. Karpyak

## Abstract

Naltrexone can aid in reducing alcohol consumption, while acamprosate supports abstinence; however, not all patients with alcohol use disorder (AUD) benefit from these treatments. Here we present the first genome-wide association study of AUD treatment outcomes based on data from the COMBINE and PREDICT studies of acamprosate and naltrexone, and the Mayo Clinic CITA study of acamprosate. Primary analyses focused on treatment outcomes regardless of pharmacological intervention and were followed by drug-stratified analyses to identify treatment-specific pharmacogenomic predictors of acamprosate and naltrexone response. Treatment outcomes were defined as: (1) time until relapse to any drinking (TR) and (2) time until relapse to heavy drinking (THR; ≥5 drinks for men, ≥4 drinks for women in a day), during the first three months of treatment. Analyses were performed within each dataset, followed by meta-analysis across the studies (N=1090 European ancestry participants). Single nucleotide polymorphisms (SNPs) in the *BRE* gene were associated with THR (min p=1.6E-08) in the entire sample, while two intergenic SNPs were associated with medication-specific outcomes (naltrexone THR: rs12749274, p=3.9E-08; acamprosate TR: rs77583603, p=3.1E-09). The top association signals for TR (p=7.7E-08) and second strongest signal in the THR (p=6.1E-08) analysis of the naltrexone-treated subset map to *PTPRD*, a gene previously implicated in addiction phenotypes in human and animal studies. Leave-one-out polygenic risk score analyses showed significant associations with TR (p=3.7E-04) and THR (p=2.6E-04). This study provides the first evidence of a polygenic effect on AUD treatment response, and identifies genetic variants associated with potentially medication-specific effects on AUD treatment response.

## INTRODUCTION

Alcohol use disorder (AUD) is highly prevalent, and presents a significant health burden worldwide [1]. Several medications are approved for treatment of AUD by the U.S. Food and Drug Administration, but they are underutilized and pharmacological treatment of AUD remains a major challenge [2]. Recent reviews have highlighted the need for a precision medicine approach in the context of AUD treatment in order to increase the utility and safety of available medications [3, 4].

Treatment efficacy of acamprosate and naltrexone is supported by large systematic meta-analyses of randomized controlled trials [5, 6], which suggest that naltrexone helps people refrain from excessive drinking while acamprosate is effective in supporting abstinence [6–12]. Despite these benefits, a considerable proportion of AUD patients fail to benefit from these drugs. The number needed to treat has been estimated to be 7-12 for acamprosate and 12-20 for naltrexone, reflecting variable response rates across studies [7, 8, 13, 14]. Garbutt et al. performed a systematic review to identify moderators of naltrexone response and concluded that available data were insufficient to guide clinical treatment [15]. Similarly, limited success was achieved in studies searching for clinical moderators of acamprosate response [16].

Although secondary analyses including application of machine learning approaches have identified potential moderators of naltrexone and acamprosate response [17], further efforts are needed to identify predictors for personalized medicine. Therefore, discovery of biomarkers of treatment response is a high priority in AUD research [18].

Genetic variation contributes to inter-individual differences in drug response [19–22], and it is expected that genomic factors can aid in prediction of response to AUD treatment [23, 24]. Moreover, discovery of genetic variation and biological pathways involved in treatment response may reveal important information about the mechanisms of drug action, accelerating further drug discovery efforts. Candidate gene studies identified several genetic variations that may influence treatment outcomes in AUD [25–28]. However, the contribution of candidate gene studies to the understanding of genetic effects on AUD treatment response has been limited, highlighting the need for genome-wide pharmacogenomic studies. Yet, to date, no genome-wide association studies (GWASs) of acamprosate or naltrexone treatment response have been published.

Recent GWASs of AUD and other alcohol use related traits have identified a growing number of loci associated with these phenotypes. Moreover, various downstream analyses, including gene and pathway-based analyses and polygenic risk score (PRS) analyses have provided further insights into the genetic architecture of AUD and related traits, including genetic overlap with other psychiatric and behavioral traits [29–32]. Analyses of quantitative measures of problem drinking and alcohol consumption in large samples have identified more genetic variations contributing to these phenotypes, while identifying loci contributing to AUD has proven more challenging due to the complexity of AUD and the difficulty of compiling large samples of patients evaluated for this phenotype and receiving similar treatments [29–32]. Despite its clinical relevance, AUD treatment response has not yet been studied with the GWAS approach. Investigating the genetics of AUD treatment response is expected to not only reveal genetic variation potentially useful in treatment selection but may also contribute to our understanding of AUD risk and prognosis.

GWAS of treatment outcomes have been performed in the context of other addictions, most notably nicotine dependence. Large studies of smoking behavior phenotypes, including smoking cessation, have demonstrated both shared and unique genetic contributions across these phenotypes, nicotine use disorder, and other diseases [33–35]. Genetic studies of nicotine clearance and nicotine metabolism biomarkers have provided other important insights regarding the genetic contributions to complex smoking related phenotypes [36, 37]. However, similar to AUD, little progress has been made in understanding pharmacogenetic factors that contribute to response to specific smoking cessation treatments.

Here we present the first GWAS of AUD treatment response based on data from three of the largest studies of acamprosate and naltrexone completed to date, with a total sample of more than 1,000 patients treated for AUD: The US-based COMBINE study [38], the German-based PREDICT study [39], and the CITA study [27]. While COMBINE and PREDICT were randomized, placebo-controlled studies designed to evaluate response to acamprosate and naltrexone, the CITA study was an open-label study of acamprosate designed for pharmacogenomics analyses. Using data from these three studies, we performed GWAS analyses of AUD treatment outcomes (across treatment options) as well as pharmacogenomics GWAS of acamprosate and naltrexone response, separately. Post-GWAS analyses were performed including gene-level tests, gene-set and tissue enrichment analyses, and polygenic risk score (PRS) analyses to evaluate polygenic predictors of treatment outcomes across datasets. Our results provide the first evidence of a polygenic signal for AUD treatment outcomes across studies.

## METHODS

### Samples

This study used a new genomic dataset derived from three previously completed studies of acamprosate and/or naltrexone treatment of AUD: the COMBINE, PREDICT, and CITA studies. The key characteristics of these three studies are summarized in Supplemental Table S1. The COMBINE study was a randomized placebo-controlled trial with 1383 alcohol dependent participants, comparing outcomes after 16 weeks of naltrexone or acamprosate, or both, with or without a combined behavioral intervention [38]. DNA for genetic studies was obtained from a subset of COMBINE study participants. PREDICT was a double-blind randomized trial comparing outcomes after three months of acamprosate, naltrexone, or placebo treatment in 426 AUD patients [39]. The CITA study was a single-arm pharmacogenomics study designed to investigate associations of acamprosate treatment outcomes with genetic polymorphisms in genes and pathways previously implicated in acamprosate response (N=443) [27]. All subjects included in our analyses provided consent allowing use of their clinical data and DNA for genetic studies of AUD and response to its treatment, and this study was approved by the Mayo Clinic Institutional Review Board.

### Genotyping, quality control and imputation

Male participants from the PREDICT study (N=266) were previously genotyped using three different Illumina® (Illumina, San Diego, CA, USA) genome-wide single nucleotide polymorphisms (SNP) platforms as part of a GWAS of alcohol dependence [40]. In CITA, samples were genotyped using Illumina® HumanCore (Illumina, San Diego, CA, USA) arrays and/or Infinium® OmniExpressExome-8 BeadChips (Illumina, San Diego, CA, USA). Of the 1383 subjects enrolled in the COMBINE study, DNA from 758 subjects was genotyped using Infinium® OmniExpressExome-8 BeadChips (Illumina, San Diego, CA, USA). All genetic data underwent quality control followed by imputation. Details of the genotyping, quality control, and imputation are included in the Supplemental Methods. After quality control, data from 498 COMBINE participants, 266 PREDICT participants and 319 CITA participants of European ancestry with treatment outcome data were available for analysis. Following imputation, 5.6 million SNPs with minor allele frequency (MAF) >0.05 in at least one of the studies were analyzed and included in the meta-analyses.

### Assessment of Treatment Outcomes

Detailed descriptions of study procedures and assessments in the COMBINE, PREDICT and CITA studies are presented elsewhere [27, 38, 39]. In brief, baseline patient characteristics were collected in each study, and outcomes were assessed during three or more months of treatment. Baseline measures included demographic data (age, sex) as well as clinical information including alcohol consumption prior to treatment initiation. Treatment outcomes were derived from timeline follow back (TLFB) data collected at different time points during a three-month period after treatment initiation. For this study, the primary treatment outcome measures were: (1) time until relapse to any drinking during the first three months of treatment, and (2) time until relapse to heavy drinking (≥5 drinks for men, ≥4 drinks for women in a day) during the first three months of treatment. Outcomes for patients that were lost to follow-up in the first three months prior to relapse (or heavy relapse) were treated as censored observations in the survival analysis. Although the two outcomes we analyzed are highly correlated because for many patients first relapse was an episode of heavy drinking, we considered both outcomes, as acamprosate is believed to be effective in preventing relapse to any drinking while naltrexone has been reported to be more effective in preventing relapse to heavy drinking [6, 8, 9].

### Genome-Wide Association Analyses

The primary analyses included patients treated with acamprosate, naltrexone, or placebo to identify predictors of treatment outcomes regardless of pharmacological intervention. Drug-stratified analyses were then run to identify treatment-specific (i.e. pharmacogenomic) predictors of acamprosate and naltrexone response. All GWAS were run in the three datasets separately, followed by fixed-effects meta-analysis. In each dataset, allelic associations with time until relapse (TR) and time until heavy relapse (THR) were assessed using Cox proportional hazards models. All analyses were adjusted for genetic principal components (PCs), if needed, to control for remaining population stratification in the European ancestry samples. The Cox proportional hazards analyses were run using the survival package in R (version 3.6.2), with SNP effect sizes estimated using hazard ratios (HR). Meta-analyses of GWAS summary statistics were performed using METAL [41].

### Gene Level Associations and Gene-Set and Tissue Enrichment Analyses

Gene-level tests were performed using Multi-marker Analysis of GenoMic Annotation (MAGMA) [42] as implemented in functional mapping and annotation of genetic associations (FUMA) [43], which uses Brown’s method to combine SNP p-values across a gene, while accounting for linkage disequilibrium (LD) between SNPs (i.e. r^2^). Competitive gene-set analyses accounting for potential confounders such as gene size and LD were also performed using MAGMA in FUMA. MAGMA implementation in FUMA was further used to perform tissue enrichment analyses that test whether association results are enriched for genes expressed in particular tissue types based on Genotype-Tissue Expression (GTEx) data [44].

### Leave-one-out Polygenic Risk Score Analyses

Leave-one-out PRS analyses were used to test for a reproducible polygenic predictor of treatment outcomes between datasets. Specifically, leave-one-out PRSs for TR and THR were generated for each target study based on the results of a discovery GWAS meta-analysis where the target cohort was left out. The PRSs were constructed using PRSice2 [45] to prune (r^2^ >0.1 within a 500 kb window) and restrict SNPs to a given p-value threshold (p_t_=0.0001, 0.001, 0.01, 0.05, 0.1, 0.2, 1), with SNP alleles weighted by their log (HR) estimates. The PRSs for TR and THR were tested for association with the respective treatment outcome using Cox proportional hazards models. Analyses of the COMBINE study data were adjusted for the first four PCs to account for the heterogeneity observed in this dataset. Finally, the results from the leave-one-out PRS analyses of the three datasets were meta-analyzed to assess overall PRS prediction of treatment response. Nagelkerke’s pseudo-R^2^ was calculated to estimate the variance explained in TR or THR by the PRS in each dataset, and weighted average R^2^ values were calculated across the three datasets using the effective N for each cohort.

## RESULTS

Descriptive statistics for the three datasets are shown in Table 1. In total, 639 (59%) of the study participants relapsed to any drinking, while 564 (52%) relapsed to heavy drinking.

**Table 1:** Demographic and Clinical Characteristics of the COMBINE, PREDICT and CITA participants included in the pharmacogenomics GWAS.

|  | COMBINE | PREDICT | CITA |
| --- | --- | --- | --- |
| Sample sizes |  |  |  |
| Total N | 498 | 266 | 319 |
| Acamprosate N | 223 | 110 | 319 |
| Naltrexone N | 199 | 102 | - |
| Placebo only or no pills N | 177 | 54 |  |
| Age, mean(SD) | 45.3 (10.6) | 45.2 (8.5) | 43.4 (11.6) |
| Sex, male N(%) | 343 (68.9%) | 266 (100%) | 204 (64.0%) |
| Age of onset | 30.5 (11.9) | 30.2 (10.2) | 29.6 (12.0) |
| Baseline alcohol consumption<sup>a</sup>, mean (sd) |  |  |  |
| Days since last drinking day | 8.0 (5.4) | 22.0 (4.4) | 23.4 (15.6) |
| Average number of drinks per drinking day | 12.7 (7.5) | 21.0 (12.7) | 11.9 (7.4) |
| % Drinking days<sup>b</sup> | 71.8 (24.8) | 82.0 (26.8) | 52.2 (29.8) |
| % heavy drinking days | 63.9 (24.8) | 79.4 (27.9) | 47.6 (30.4) |
| Treatment Outcomes: |  |  |  |
| Relapse, N (%) | 380 (76.3%) | 158 (59.4%) | 101 (31.7%) |
| Relapse in acamprosate subset, N (%) | 167 (74.9%) | 62 (56.4%) | 101 (31.7%) |
| Relapse in naltrexone subset, N (%) | 146 (73.4%) | 65 (63.7%) | - |
| Heavy Relapse, N (%) | 338 (67.9%) | 142 (53.4%) | 84 (26.3%) |
| Heavy relapse in acamprosate subset, N (%) | 144 (64.6%) | 56 (50.9%) | 84 (26.3%) |
| Heavy relapse in naltrexone subset, N (%) | 127 (63.8%) | 57 (55.9%) | - |

<sup>a</sup>Baseline alcohol consumption measures are based on 90 days before start of treatment in COMBINE and CITA, and 30 days before start of treatment in PREDICT.
<sup>b</sup>% drinking days = 100 - % days abstinent in the same time period

### Genome-wide association study

Manhattan plots for the two GWAS meta-analyses in the full cohort (all treatments) are shown in Figure 1; the corresponding Quantile/Quantile (QQ) plots are shown in Supplemental Figure S1. In the meta-analysis across the three studies no individual SNPs were significantly associated with TR; the SNP with the strongest evidence for association is located in an intron of *KCNQ4* (rs1078110, P=6.2E-07). In the full cohort analysis of THR, significant associations were observed for a set of SNPs in *BRE* (14 genome-wide significant SNPs, top SNP rs56951679, minor allele HR=1.5, p=1.6E-08). The effect estimate for the top SNP at this locus (rs56951679) suggests that carrying one additional copy of the minor allele (C) is associated with 1.5 times higher risk of relapse to heavy drinking. A regional association plot for this locus is shown in Supplemental Figure S2.

**Figure 1:**
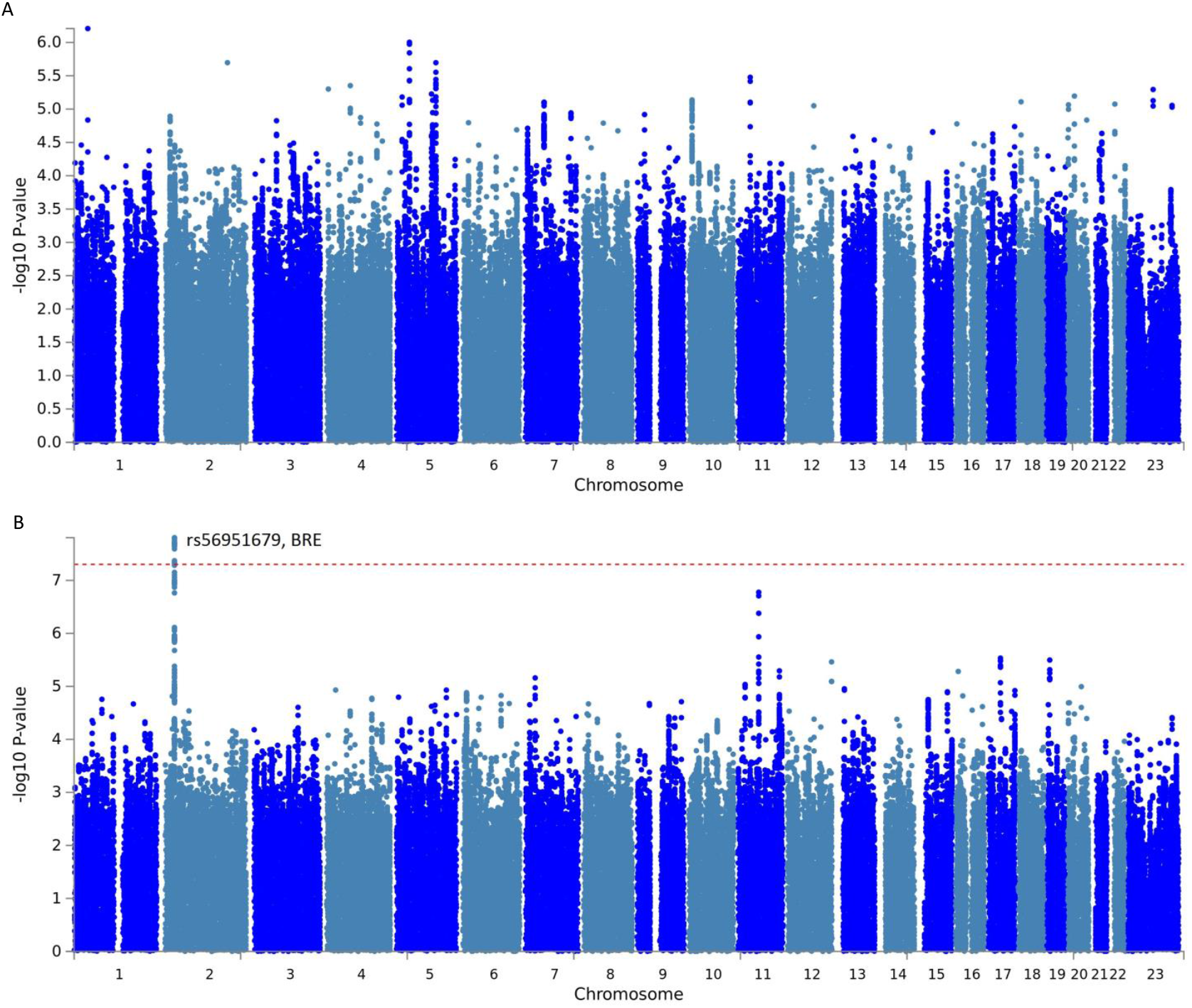
Manhattan Plots for (A) Time until relapse to any drinking and (B) Time until relapse to heavy drinking, in all subjects.

In drug-stratified meta-analyses of outcomes in patients treated with naltrexone (Supplemental Figure S3), an intergenic SNP located between the long intergenic non-coding RNA RP4-710M16.2 and the *PPAP2B* gene was associated with THR (rs12749274, HR=2.90, p=3.9E-08). While not achieving genome-wide significance, the strongest evidence for association with TR and second strongest for THR was observed with a SNP in an intron of *PTPRD* (rs62533259: TR minor allele HR=2.2, p=7.7E-08; THR minor allele HR=2.4, p=6.1E-08). *PTPRD* has been implicated in multiple addiction related phenotypes in humans and other species[46].

One SNP was significantly associated with TR during acamprosate treatment; this intergenic SNP is located between a non-coding RNA gene and the ribosomal protein L29 pseudogene (rs77583603; p=3.1E-09; Supplemental Figure S4). The top signal in the analyses of THR in acamprosate-treated patients was rs34797278 in a non-coding RNA (p=5.4E-08). All loci with suggestive evidence of association (p<5*10^−6^) with one of the outcome measures are shown in Supplementary Tables S1 (all treatments), S2 (naltrexone), and S3 (acamprosate).

Supplementary Figure S5 shows scatterplots of results (p-values) from analyses of TR vs. THR, and for analyses of acamprosate treatment outcomes vs. naltrexone treatment outcomes. As expected, TR GWAS signals were highly correlated with THR signals (panels A, B, and C of Supplementary Figure S5), but results for acamprosate treatment outcomes showed little correlation with signals for naltrexone treatment outcomes (panels D and E of Supplementary Figure S5). Although TR signals were generally highly correlated with THR signal, the top association from the THR analysis of the full cohort (the *BRE* top SNP p=1.6E-08) showed much weaker evidence for association with TR (p=1E-04). On the other hand, the *PTPRD* rs62533259 association in the naltrexone-treated subset was a top signal in both the TR and THR analyses (shown in panel B of Supplemental Figure S5). However, the same *PTPRD* variants were not associated with TR or THR in the acamprosate-treated patient subset (noted in panels D and E of Supplemental Figure S5).

### Gene-level, Gene-Set, and Tissue Enrichment Analyses

There were no significant gene-level associations based on MAGMA analyses (see Methods for details) of the GWAS results presented above, although the *BRE* gene association with THR in the full sample approached significance (p=4.0E-6, Bonferroni-corrected p=0.079). In MAGMA gene-set association analyses of THR in naltrexone-treated patients, the strongest association was with a curated gene set named “boyault_liver_cancer_subclass_g56_up” representing up-regulated genes in hepatocellular carcinoma subclass G56, defined by unsupervised clustering; the 11 genes in this gene set showed enrichment for association with THR in the naltrexone-treated patients (p=1.0E-06, Bonferroni corrected p=0.016; most strongly associated genes in this gene set were *DPP4, SMYD2*, and *TBX3*). Also, in the analyses of naltrexone treatment outcomes, the top tissues based on expression of genes enriched for association signals (at p<0.05) were brain tissues, including the hippocampus, putamen, basal ganglia, cortex, and amygdala (Supplemental Fig S6), but these enrichment results were not statistically significant after multiple testing correction.

### Leave-one-out Polygenic Prediction

Leave-one-out analyses provided evidence of a polygenic effect on treatment response in AUD. Specifically, for both TR and THR, PRSs derived from SNP effects observed in any two of the studies (using p-value threshold of 0.05) explained a statistically significant proportion of variance in the same outcome in the left-out dataset (Nagelkerke’s R^2^= 1.3%; p=3.7E-04 for time until relapse, Nagelkerke’s R^2^=1.3%; p=2.6E-04 for time until heavy relapse; Figure 2).

**Figure 2:**
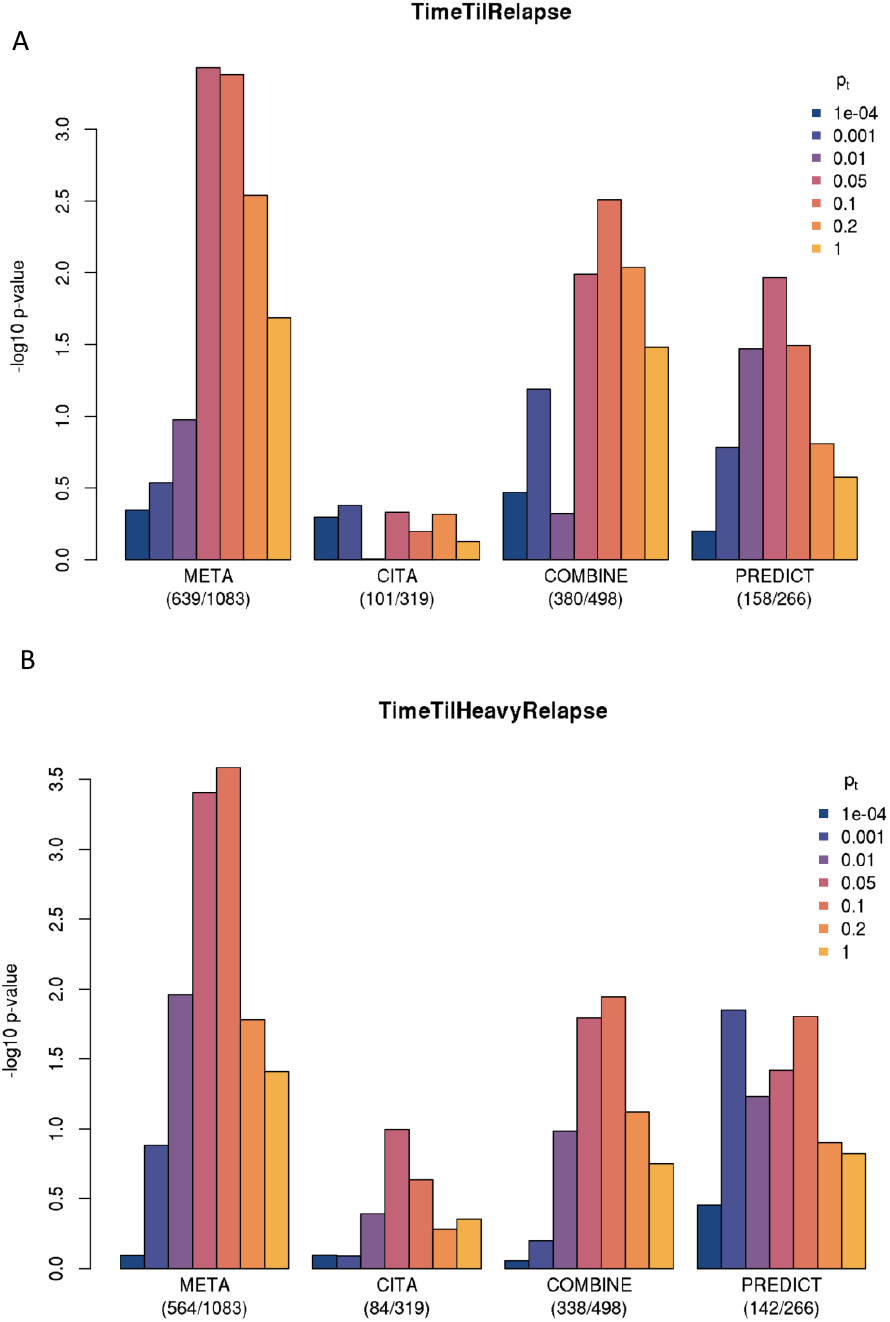
Leave-one-out PRS analysis of (A) time until relapse to any drinking and (B) time until relapse to heavy drinking. In each of the three studies (CITA, COMBINE and PREDICT) PRSs were constructed based on a discovery GWAS in the remaining two samples across a range of p-value thresholds (p_T_ denoted using different colors described in the legend) to select SNPs for inclusion in the PRS. The selected SNPs (after LD pruning) were used to compute PRSs and the association of the PRSs with the outcome (TR or THR) was tested. The plots show the –log10(p-values) for these association tests (on the y-axis) in each sample, as well as the meta-analysis of the leave-one-out PRS associations across the studies. The PRS association meta-analyses provided significant results for both time until relapse (p=3.7E-04, Nagelkerke’s R^2^ = 1.3% at pT=0.05) and time until heavy relapse (p=2.6E-04, Nagelkerke’s R^2^ = 1.3% at p_T_ =0.10).

## DISCUSSION

Using data from three studies of AUD treatment outcomes, we performed the first GWAS for AUD treatment response, including drug-stratified pharmacogenomic analyses of acamprosate and naltrexone treatment outcomes, and we identified several genome-wide significant associations. We also performed PRS analyses to assess if a polygenic signal captured by this GWAS is associated with AUD treatment outcomes in independent samples. Results of these leave-one-out PRS analyses provided the first evidence of association between AUD treatment outcomes and PRSs, reflecting combined effect of variation across the genome on response to AUD treatment. While the proportion of variation in the treatment response explained by the PRSs is small, as expected for relatively small studies of complex traits, the significant association demonstrates that polygenic effects contribute to AUD treatment outcomes motivating further research into identifying contributing genetic factors.

Significant evidence of association was observed between THR and SNPs in the brain and reproductive organ-expressed protein (*BRE*) gene (aliases include *BABAM2, BRCC4* and *BRCC45*), which encodes BRISC and BRACA1 A complex member 2. *BRE* is ubiquitously expressed in human tissues, but most prominently in the zona glomerulosa of the adrenal cortex, where mineralocorticoids (e.g., aldosterone) are synthesized and secreted, as well as in glia and neurons [47]. Although the exact role of *BRE* in mineralocorticoid synthesis and secretion is unclear, *BRE* expression is altered in adrenal abnormalities, suggesting *BRE* involvement in adrenal function [47]. Chronic alcohol consumption has been shown to increase blood aldosterone levels in humans and other species, and increased aldosterone level was correlated with higher alcohol consumption, craving and anxiety levels in AUD patients [48].

Aldosterone may regulate alcohol use behaviors and anxiety via mineralocorticoid receptors expressed in limbic regions [49], including regions involved in regulating anxiety, stress-induced alcohol consumption, craving, and inhibitory control. Thus, our finding of association of *BRE* SNPs with THR may be related to regulation of stress response and alcohol craving.

Furthermore, *BRE* has shown genome-wide significant associations with a range of phenotypes from different domains (e.g. metabolic, immunological, cardiovascular) [50]. With respect to psychiatric traits, genetic variation in *BRE* has been associated with alcohol intake frequency and drinks per week in the general population [33]. However, it is also worth noting that according to GTEx, *BRE* rs56951679 (and other top SNPs in our analysis) is a splicing qualitative trait locus (sQTL) for *FNDC4* exon 7 in human cortex, hippocampus and nucleus accumbens. FNDC4, a secreted factor highly expressed in liver and brain, was implicated in sex-specific neurotoxic consequences of chronic alcohol withdrawal [51]. The mechanisms of action through which variants in the *BRE* region may impact response to AUD treatment need to be further explored.

The one SNP with genome-wide significant evidence of association with acamprosate treatment response (rs77583603; p=3.1E-09 for TR) is located between a non-coding RNA gene and the ribosomal protein L29 pseudogene and is not itself annotated as functionally significant.

Analyses of data from patients treated with naltrexone identified a SNP near *PPAP2B* that was significantly associated with THR. *PPAP2B* (a.k.a. *PLPP3, LPP3*) encodes phospholipid phosphatase 3. Changes in *PPAP2B* gene expression have been observed in nucleus accumbens and amygdala of rodents after voluntary ethanol consumption [52, 53], and *PPAP2B* has been implicated in alcoholic fatty liver in humans [54]. These genes/variants identified in the medication-specific analyses have not been previously implicated in AUD or other addiction-related traits in humans, and their potential role in AUD treatment response needs further investigation.

An intronic SNP of *PTPRD*, while not quite genome-wide significant, was the strongest association signal for TR (p=7.7E-08) and the second strongest association for THR (p=6.1E-08) in the naltrexone analysis. This finding is notable because *PTPRD* (protein tyrosine phosphatase receptor type D) has been implicated in multiple addiction related phenotypes in humans and in animal studies [55, 56]. Protein tyrosine phosphatases are signaling molecules that regulate a variety of cellular processes, and *PTPRD* likely plays a role in neuronal cell adhesion. Uhl and Martinez [46] provided a comprehensive review of *PTPRD* genetics and neurobiology, and discussed its potential role as a pharmacological target with effects on brain phenotypes. Of note, the *PTPRD* effect we observed appears to be a pharmacogenomic effect specific to naltrexone, as our study provided no evidence of association of the same *PTPRD* variant with acamprosate treatment response. The impact of *PTPRD* on naltrexone treatment outcomes is, therefore, intriguing and warrants further investigation.

We performed GWAS of treatment outcomes in the full cohort of patients enrolled in the COMBINE, PREDICT and CITA studies, as well as drug-specific analyses of treatment outcomes for acamprosate and naltrexone. The drug-stratified analyses aim to identify genetic variants that impact response to a given drug. The analyses of the full sample may also identify pharmacogenomic effects, but have greater power to identify genetic variants that impact risk of relapse regardless of specific treatment – these variants may include markers of severity of AUD or AUD subtypes that confer differential risk of relapse following treatment. Two genes implicated in this study, *BRE* and *PTPRD*, were previously associated with AUD or related phenotypes. Several known AUD risk genes, particularly *ADH1B*, that have been most consistently associated with AUD in large studies [32], were not significant predictors of treatment response in our analyses. The lack of such findings may be partly due to a relatively small contribution of AUD risk genes to treatment response, which would be consistent with findings in other psychiatric disorders including major depressive disorder [57, 58]. Additionally, low-frequency variants that are protective against AUD could have very low frequencies in the studied samples of AUD patients, reducing the power to detect their contribution to treatment response. Indeed, this may be the case with *ADH1B* variants that are protective against AUD but have low allele frequencies in populations of European ancestry. For example, the well-established AUD-associated SNP rs1229984 in *ADH1B* has a frequency of ∼5% in European populations [59]. In the COMBINE dataset this SNP had a frequency of only ∼2%, whereas in CITA it had a frequency of ∼1%, and in PREDICT its frequency was <1%. Because of its low frequency and poor imputation quality in the CITA and PREDICT samples, this SNP was not analyzed in these datasets. However, analysis of the COMBINE dataset provided marginal evidence of association for rs1229984 with TR (HR=0.32, p=0.016) and THR (HR=0.27, p=0.0053); the effect estimates suggest the minor allele at rs1229984, which is protective against AUD, may have a strong protective effect against relapse during treatment, reducing the risk of relapse by about 70%. Because its MAF was below 0.05 in all of the samples, this SNP was excluded from the meta-analyses. As evidence accumulates regarding other genetic variants that contribute to AUD and related phenotypes, and as larger studies allow the computation of more predictive polygenic risk scores for AUD and related traits, it will be important to further examine the association of those variants and polygenic risks with AUD treatment outcomes.

This study made use of data from three prior studies of AUD treatment. These three studies have important similarities allowing for combined analysis of treatment outcomes, including using the TLFB to assess drinking outcomes. However, the three studies also differed in relevant patient characteristics. The available PREDICT study data were only from men, but the COMBINE and CITA study samples included 31% and 36% women, respectively. Reported pre-treatment baseline alcohol consumption also differed, with PREDICT participants having the highest (and CITA participants having the lowest) alcohol consumption at baseline and participants in the COMBINE study having the shortest duration of abstinence prior to treatment initiation. The studies also differed in rates of treatment outcomes, with the CITA sample having the lowest percentage of any-drinking relapse and heavy-drinking relapse. While this heterogeneity may have limited the replicability of findings between the three datasets and reduced power of the meta-analysis, the differences between the studies also mean that identified genetic associations are likely robust genetic predictors of treatment outcomes that apply to a broad range of patients.

In addition to sample heterogeneity that may have reduced the power of this study, other limitations need to be noted. The sample size of this study is smaller than what is typically required for well-powered GWAS of complex traits, particularly when the goal is identification of specific genetic risk variants. However, this sample size of approximately 1,000 was expected to be adequate to detect polygenic effects, as supported by our PRS results. Other limitations of this study include a lack of thorough investigation of the role of intermediate or potentially confounding factors such as demographic and clinical variables or comorbidities (e.g. age, sex, smoking) as well as measures of AUD severity or alcohol consumptions. Because each of the contributing studies collected different data at baseline, harmonization of these variables and investigation of their role in the context of pharmacogenomics effects on AUD treatment response was beyond the scope of this study, but should be further investigated in the future. Finally, because of the limited ancestral diversity in the contributing studies, the analysis was limited to patients of European ancestry, limiting the generalizability of the findings to other populations, and reducing the power to identify effects of variants that are rare in European ancestry populations such as the *ADH1B* variant discussed above, or the *ALDH2* Glu504Lys polymorphism (rs671) that generates extraordinarily significant association to AUD (e.g. p = 10^−4740^), but only in populations where the locus is actually polymorphic. Studies of the genetics of AUD treatment response in samples with other ancestries are needed.

In conclusion, these first GWAS analyses of AUD treatment outcomes had limited power for discovery of specific genetic variants associated with response to AUD treatment. Nevertheless, they provided important insights, including the first demonstration of polygenic effects on AUD treatment outcomes, and identification of genetic variants potentially associated with AUD treatment response. These findings motivate further investigation of the genetic contribution to AUD outcomes and study of the biological mechanisms underlying response to medications used in the treatment of AUD.

## Data Availability

Data is not able to be shared without approved project proposal to individual studies.

## Acknowledgements

This work was supported by the National Institute on Alcohol Abuse and Alcoholism grants R21 AA25214, U01 AA027487, and R01 AA27486, and by the German Federal Ministry of Education and Research (BMBF) through grant SysmedSUD 01ZX01909A. Collection of data for the CITA study was supported by P20 AA017830.

## References

1. Alcohol, G.B.D. and C. Drug Use, The global burden of disease attributable to alcohol and drug use in 195 countries and territories, 1990-2016: a systematic analysis for the Global Burden of Disease Study 2016. Lancet Psychiatry, 2018. 5(12): p. 987–1012.

2. Litten, R.Z., et al., Discovery, Development, and Adoption of Medications to Treat Alcohol Use Disorder: Goals for the Phases of Medications Development. Alcohol Clin Exp Res, 2016. 40(7): p. 1368–79.

3. Hartwell, E.E. and H.R. Kranzler, Pharmacogenetics of alcohol use disorder treatments: an update. Expert Opin Drug Metab Toxicol, 2019. 15(7): p. 553–564.

4. Helton, S.G. and F.W. Lohoff, Pharmacogenetics of alcohol use disorders and comorbid psychiatric disorders. Psychiatry Res, 2015. 230(2): p. 121–9.

5. Donoghue, K., et al., The efficacy of acamprosate and naltrexone in the treatment of alcohol dependence, Europe versus the Rest of the World: a meta-analysis. Addiction, 2015.

6. Jonas, D.E., et al., Pharmacotherapy for adults with alcohol use disorders in outpatient settings: a systematic review and meta-analysis. JAMA : the journal of the American Medical Association, 2014. 311(18): p. 1889–900.

7. Bouza, C., et al., Efficacy and safety of naltrexone and acamprosate in the treatment of alcohol dependence: a systematic review. Addiction, 2004. 99(7): p. 811–28.

8. Rosner, S., et al., Acamprosate supports abstinence, naltrexone prevents excessive drinking: evidence from a meta-analysis with unreported outcomes. Journal of Psychopharmacology, 2008. 22(1): p. 11–23.

9. Maisel, N.C., et al., Meta-analysis of naltrexone and acamprosate for treating alcohol use disorders: when are these medications most helpful? Addiction, 2013. 108(2): p. 275–93.

10. Dawson, D.A., R.B. Goldstein, and B.F. Grant, Rates and correlates of relapse among individuals in remission from DSM-IV alcohol dependence: a 3-year follow-up. Alcoholism, Clinical and Experimental Research, 2007. 31(12): p. 2036–45.

11. Witkiewitz, K., K. Saville, and K. Hamreus, Acamprosate for treatment of alcohol dependence: mechanisms, efficacy, and clinical utility. Therapeutics and clinical risk management, 2012. 8: p. 45–53.

12. Cheng, H.Y., et al., Treatment interventions to maintain abstinence from alcohol in primary care: systematic review and network meta-analysis. BMJ, 2020. 371: p. m3934.

13. Rosner, S., et al., Acamprosate for alcohol dependence. The Cochrane database of systematic reviews, 2010(9): p. CD004332.

14. Heilig, M. and M. Egli, Pharmacological treatment of alcohol dependence: target symptoms and target mechanisms. Pharmacology and Therapeutics, 2006. 111(3): p. 855–76.

15. Garbutt, J.C., et al., Clinical and biological moderators of response to naltrexone in alcohol dependence: a systematic review of the evidence. Addiction, 2014. 109(8): p. 1274–84.

16. Verheul, R., et al., Predictors of acamprosate efficacy: results from a pooled analysis of seven European trials including 1485 alcohol-dependent patients. Psychopharmacology, 2005. 178(2-3): p. 167–73.

17. Gueorguieva, R., et al., Predictors of abstinence from heavy drinking during treatment in COMBINE and external validation in PREDICT. Alcoholism, Clinical and Experimental Research, 2014. 38(10): p. 2647–56.

18. Litten, R.Z., et al., Five Priority Areas for Improving Medications Development for Alcohol Use Disorder and Promoting Their Routine Use in Clinical Practice. Alcohol Clin Exp Res, 2020. 44(1): p. 23–35.

19. Wilson, J.F., et al., Population genetic structure of variable drug response. Nature Genetics, 2001. 29(3): p. 265–9.

20. Weinshilboum, R.M. and L. Wang, Pharmacogenetics and pharmacogenomics: development, science, and translation. Annual review of genomics and human genetics, 2006. 7: p. 223–45.

21. Wong, M.L., et al., Prediction of susceptibility to major depression by a model of interactions of multiple functional genetic variants and environmental factors. Molecular Psychiatry, 2012. 17(6): p. 624–33.

22. van der Wouden, C.H., et al., Development of the PGx-Passport: A Panel of Actionable Germline Genetic Variants for Pre-Emptive Pharmacogenetic Testing. Clin Pharmacol Ther, 2019. 106(4): p. 866–873.

23. Kranzler, H.R. and H.J. Edenberg, Pharmacogenetics of alcohol and alcohol dependence treatment. Current Pharmaceutical Design, 2010. 16(19): p. 2141–8.

24. Litten, R.Z., A.M. Bradley, and H.B. Moss, Alcohol biomarkers in applied settings: recent advances and future research opportunities. Alcoholism, Clinical and Experimental Research, 2010. 34(6): p. 955–67.

25. Anton, R.F., et al., An evaluation of mu-opioid receptor (OPRM1) as a predictor of naltrexone response in the treatment of alcohol dependence: results from the Combined Pharmacotherapies and Behavioral Interventions for Alcohol Dependence (COMBINE) study. Archives of General Psychiatry, 2008. 65(2): p. 135–44.

26. Kiefer, F., et al., Involvement of the atrial natriuretic peptide transcription factor GATA4 in alcohol dependence, relapse risk and treatment response to acamprosate. The pharmacogenomics journal, 2011. 11(5): p. 368–74.

27. Karpyak, V.M., et al., Genetic markers associated with abstinence length in alcohol-dependent subjects treated with acamprosate. Translational psychiatry, 2014. 4: p. e453.

28. Spanagel, R., et al., The clock gene Per2 influences the glutamatergic system and modulates alcohol consumption. Nature Medicine, 2005. 11(1): p. 35–42.

29. Clarke, T.K., et al., Genome-wide association study of alcohol consumption and genetic overlap with other health-related traits in UK Biobank (N=112 117). Mol Psychiatry, 2017. 22(10): p. 1376–1384.

30. Deak, J.D., A.P. Miller, and I.R. Gizer, Genetics of alcohol use disorder: a review. Curr Opin Psychol, 2019. 27: p. 56–61.

31. Kranzler, H.R., et al., Genome-wide association study of alcohol consumption and use disorder in 274,424 individuals from multiple populations. Nat Commun, 2019. 10(1): p. 1499.

32. Walters, R.K., et al., Transancestral GWAS of alcohol dependence reveals common genetic underpinnings with psychiatric disorders. Nat Neurosci, 2018. 21(12): p. 1656–1669.

33. Liu, M., et al., Association studies of up to 1.2 million individuals yield new insights into the genetic etiology of tobacco and alcohol use. Nat Genet, 2019. 51(2): p. 237–244.

34. Matoba, N., et al., GWAS of smoking behaviour in 165,436 Japanese people reveals seven new loci and shared genetic architecture. Nat Hum Behav, 2019. 3(5): p. 471–477.

35. Saccone, N.L., et al., Genome-Wide Association Study of Heavy Smoking and Daily/Nondaily Smoking in the Hispanic Community Health Study/Study of Latinos (HCHS/SOL). Nicotine Tob Res, 2018. 20(4): p. 448–457.

36. Buchwald, J., et al., Genome-wide association meta-analysis of nicotine metabolism and cigarette consumption measures in smokers of European descent. Mol Psychiatry, 2020.

37. Chenoweth, M.J., et al., Genome-wide association study of a nicotine metabolism biomarker in African American smokers: impact of chromosome 19 genetic influences. Addiction, 2018. 113(3): p. 509–523.

38. Anton, R.F., et al., Combined pharmacotherapies and behavioral interventions for alcohol dependence: the COMBINE study: a randomized controlled trial. JAMA : the journal of the American Medical Association, 2006. 295(17): p. 2003–17.

39. Mann, K., et al., Searching for responders to acamprosate and naltrexone in alcoholism treatment: rationale and design of the PREDICT study. Alcoholism, Clinical and Experimental Research, 2009. 33(4): p. 674–83.

40. Treutlein, J., et al., Genome-wide association study of alcohol dependence. Arch Gen Psychiatry, 2009. 66(7): p. 773–84.

41. Willer, C.J., Y. Li, and G.R. Abecasis, METAL: fast and efficient meta-analysis of genomewide association scans. Bioinformatics, 2010. 26(17): p. 2190–1.

42. de Leeuw, C.A., et al., MAGMA: generalized gene-set analysis of GWAS data. PLoS Comput Biol, 2015. 11(4): p. e1004219.

43. Watanabe, K., et al., Functional mapping and annotation of genetic associations with FUMA. Nat Commun, 2017. 8(1): p. 1826.

44. Consortium, G.T., The Genotype-Tissue Expression (GTEx) project. Nat Genet, 2013. 45(6): p. 580–5.

45. Choi, S.W. and P.F. O’Reilly, PRSice-2: Polygenic Risk Score software for biobank-scale data. Gigascience, 2019. 8(7).

46. Uhl, G.R. and M.J. Martinez, PTPRD: neurobiology, genetics, and initial pharmacology of a pleiotropic contributor to brain phenotypes. Ann N Y Acad Sci, 2019. 1451(1): p. 112–129.

47. Miao, J., et al., Differential expression of a stress-modulating gene, BRE, in the adrenal gland, in adrenal neoplasia, and in abnormal adrenal tissues. J Histochem Cytochem, 2001. 49(4): p. 491–500.

48. Aoun, E.G., et al., A relationship between the aldosterone-mineralocorticoid receptor pathway and alcohol drinking: preliminary translational findings across rats, monkeys and humans. Mol Psychiatry, 2018. 23(6): p. 1466–1473.

49. Koning, A., et al., Glucocorticoid and Mineralocorticoid Receptors in the Brain: A Transcriptional Perspective. J Endocr Soc, 2019. 3(10): p. 1917–1930.

50. Watanabe, K., et al., A global overview of pleiotropy and genetic architecture in complex traits. Nat Genet, 2019. 51(9): p. 1339–1348.

51. Hashimoto, J.G. and K.M. Wiren, Neurotoxic consequences of chronic alcohol withdrawal: expression profiling reveals importance of gender over withdrawal severity. Neuropsychopharmacology, 2008. 33(5): p. 1084–96.

52. Lesscher, H.M., et al., Amygdala 14-3-3ζ as a novel modulator of escalating alcohol intake in mice. PLoS One, 2012. 7(5): p. e37999.

53. Bell, R.L., et al., Gene expression changes in the nucleus accumbens of alcohol-preferring rats following chronic ethanol consumption. Pharmacol Biochem Behav, 2009. 94(1): p. 131–47.

54. Covarrubias, M.Y., et al., Chronic alcohol exposure alters transcription broadly in a key integrative brain nucleus for homeostasis: the nucleus tractus solitarius. Physiol Genomics, 2005. 24(1): p. 45–58.

55. Drgonova, J., et al., Mouse Model for Protein Tyrosine Phosphatase D (PTPRD) Associations with Restless Leg Syndrome or Willis-Ekbom Disease and Addiction: Reduced Expression Alters Locomotion, Sleep Behaviors and Cocaine-Conditioned Place Preference. Mol Med, 2015. 21(1): p. 717–725.

56. Uhl, G.R., et al., Cocaine reward is reduced by decreased expression of receptor-type protein tyrosine phosphatase D (PTPRD) and by a novel PTPRD antagonist. Proc Natl Acad Sci U S A, 2018. 115(45): p. 11597–11602.

57. Garcia-Gonzalez, J., et al., Pharmacogenetics of antidepressant response: A polygenic approach. Prog Neuropsychopharmacol Biol Psychiatry, 2017. 75: p. 128–134.

58. Ward, J., et al., Polygenic risk scores for major depressive disorder and neuroticism as predictors of antidepressant response: Meta-analysis of three treatment cohorts. PLoS One, 2018. 13(9): p. e0203896.

59. Lek, M., et al., Analysis of protein-coding genetic variation in 60,706 humans. Nature, 2016. 536(7616): p. 285–91.

