## SupplementaryMaterial for "Genetic Contributions to Alcohol Use Disorder Treatment Outcomes: A Genome-wide Pharmacogenomics Study"

Contents:

Supplemental Methods: Genotyping, QC and Imputation

Supplemental Table S1: Characteristics of the three contributing AUD treatment studies

Supplemental Table S2: Loci* with suggestive evidence of association (p<5E-06) with one of the outcome measures when analyzing the full cohort (all treatments)

Supplemental Table S3: Loci with suggestive evidence of association (p<5E-06) with one of the outcome measures in analyses limited to patients treated with naltrexone

Supplemental Table S4: Loci with suggestive evidence of association (p<5E-06) with one of the outcome measures in analyses limited to patients treated with acamprosate

Supplemental Figure S1: QQ plots for time until relapse and time until heavy relapse in the analysis of data from all patients

Supplemental Figure S2: Regional association plot for the BRE locus in the full cohort analysis of time until heavy relapse.

Supplemental Figure S3: Manhattan and QQ plots for time until relapse and time until heavy relapse in naltrexone-treated patients

Supplemental Figure S4: Manhattan and QQ plots for time until relapse and time until heavy relapse in acamprosate-treated patients

Supplemental Figure S5: Scatterplots comparing results (p-values) from GWAS of TR vs. THR, and comparing results of analyses of different patient subsets.

Supplemental Figure S6: MAGMA Tissue Expression Analysis of THR in the naltrexone-treated patients.

**Supplemental Methods: Genotyping, Quality Control (QC), and Imputation**

Male participants from the PREDICT study were previously genotyped using Illumina HumanHap550v3, Illumina human610, or Illumina Human 660quad genotyping chips as part of a GWAS of alcohol dependence [1]. Genetic data for SNPs present on all three platforms underwent QC, as previously described [1, 2]. Data for N=266 subjects and ~464k SNPs passed QC, and were available for our pharmacogenomics analyses.

CITA samples were first genotyped using Illumina HumanCore (N=433) arrays at the Medical Genome Facility at Mayo Clinic, and subsequently were re-genotyped using the denser Infinium OmniExpressExome-8 BeadChips (N=437, including N=400 previously genotyped samples) at National Institute on Alcohol Abuse and Alcoholism (NIAAA) Laboratory of Neurogenetics. Data from the two arrays were quality-controlled, combined and checked for concordance, and additional QC was performed on the combined dataset. Samples were excluded from analysis if they had a low call rate, extreme heterozygosity, or disagreement between reported sex and genetically determined sex. Sample relatedness was checked by pairwise identical-by-descent (IBD) estimation and one subject was removed from each pair with proportion IBD (i.e. PI_HAT) >.2. SNPs were excluded from analysis if they had a call rate <99%, or deviated significantly from Hardy Weinberg Equilibrium (p<1E-06). Genetic data from 436 subjects passed QC, all with call rates >.995 and >70% European ancestry based on STRUCTURE [3] analysis with the 1000 genomes data as a reference sample were retained for analysis.

Of the 1383 subjects enrolled in the COMBINE study, DNA from 758 subjects was genotyped using Infinium OmniExpressExome-8 BeadChips at NIAAA Laboratory of Neurogenetics, and Standard QC was performed similarly to the CITA sample (excluding SNPs and samples with call rates <.95, SNPs with HWE p<1E-06, samples with discrepancies between reported sex and genetically determined sex, and one sample from each pair with IBD PI_HAT>.2). Additional QC filters were then applied, including genotype concordance checks with a prior candidate gene panel to identify potential sample mismatches. After QC, 505 subjects with >75% European ancestry based on STRUCTURE [3] analysis with the 1000 genomes data as a reference sample were retained for analysis.

Analyses were restricted to subjects of European ancestry. Imputation was performed separately for each study (436 CITA, 266 PREDICT and 505 COMBINE subjects of European ancestry) using the Michigan Imputation Server with the HRC reference panel (version HRC.r1-1.GRCh37.wgs.mac5.sites). For each dataset, variants with imputation dosage R^2^ <.5 and MAF < 0.01 were excluded from analysis.

**Supplementary Table S1:** Characteristics of the three contributing AUD treatment studies.

|  | **COMBINE** | **PREDICT** | **CITA** |
| --- | --- | --- | --- |
| Study design | Double-blind randomized controlled trial  (Non-medication interventions were not blinded to participants) | Double-blind randomized controlled trial | Open-label uncontrolled trial  (Designed to identify genetic markers associated with treatment response) |
| Study treatment | Nine arms:   1. Medical Management + Medication:    1.1. Naltrexone & acamprosate    1.2. Naltrexone    1.3. Acamprosate    1.4. Placebo 2. Medical Management + Medication + COMBINE Behavioral Intervention (CBI):    2.1. Naltrexone & acamprosate    2.2. Naltrexone    2.3. Acamprosate    2.4. Placebo 3. CBI only (no pills)   *Dose: Naltrexone (2×50 mg/d)  *Dose: Acamprosate (3×2tablets×500mg) | Three arms:   1. Naltrexone (1×50 mg/d) 2. Acamprosate (3×2tablets×333mg) 3. Placebo   *With biweekly Medical Management, i.e., manualized supportive therapy (until week 24)  *If relapsed, subjects were offered an inpatient intervention followed by re-randomization to either Medical Management or Cognitive Behavioral Intervention | One arm:   1. Acamprosate (3×2tablets×333mg)   *Attendance and involvement of Alcoholics Anonymous were encouraged and self-monitored |
| Treatment received at recruitment | Various (7.7% received inpatient detoxification service within 30 days prior to randomization) | Inpatients detoxification program | Inpatients detoxification program or outpatient program  *Comorbid depression and/or anxiety disorders were treated with antidepressants when necessary |
| Drug treatment trial period | 16 weeks | 12 weeks | 12 weeks |
| Medication compliance check | Self-report based on timeline-follow back, pill count, and confirmatory plasma levels | Self-report and pill count | Self-report and pill count |
| Follow-up schedules | Medical Management: weeks 0, 1, 2, 4, 6, 8, 10, 12, and 16  CBI: up to 20 sessions  In-person visits: weeks 8, 16, 26, 52, and 68 | Biweekly for Medical Management, then in-person visits: months 9, 12, 15, and 18 | In-person visits: months 1, 3, and 6  Phone calls: months 2, 4, and 5 |

| **(Cont’d)** | **COMBINE** | **PREDICT** | **CITA** |
| --- | --- | --- | --- |
| Sites of recruitment | U.S.A.: 11 academic sites | Germany: 5 Academic/University health centers and 2 psychiatric state hospitals | U.S.A.: Academic medical center (Mayo Clinic at Rochester, and 3 Mayo Health System sites) |
| Inclusion criteria | - Males or females age ≥ 21 years - Current DSM-IV diagnosis of alcohol dependence - Minimum of 14 drinks (females) or 21 drinks (males) on average per week over a consecutive 30-day period AND ≥2 days of heavy drinking (4 drinks for females, 5 drinks for males) within the 90 days prior to initiation of abstinence - At least 72 hours of abstinence and no significant withdrawal symptoms (CIWA<8) prior to randomization - Maximum of 21 days of abstinence prior to randomization - ≤ 21 consecutive days of planned absence during the 6-month active treatment period - A witnessed declaration of informed consent signed | - Males or females age >18 but < 65 years - Current DSM-IV/ICD-10 diagnosis of alcohol dependence - Minimum of 14 drinks (females) or 21 drinks (males) on average per week over a consecutive 30-day period AND ≥2 days of heavy drinking (4 drinks for females, 5 drinks for males) within the 90 days prior to initiation of abstinence - At least 72 hours of abstinence and no significant withdrawal symptoms (CIWA<8) prior to randomization - At least 2 weeks of inpatient detoxification - Maximum of 28 days of abstinence prior to randomization - Agree not to seek additional psychotherapy during the first 6 months of study (except mutual help groups) - A witnessed declaration of informed consent signed | - Male or females age 18-80 years - Current primary diagnosis of alcohol dependence based on DSM-IV-TR criteria - Last drink at least 5 days but not more than 6 months prior to enrollment - Enrollment in the IRB approved protocol “Developing a DNA Repository for Genomic Studies of Addiction” |
| Exclusion criteria | - Concurrent DSM-IV criteria for depression, bipolar disorder, schizophrenia, bulimia/anorexia, dementia, or a psychological disorder for whom medication is indicated (but not other Axis I disorders that are unmedicated) - Intend to engage with concurrent psychiatric treatment for alcohol-related problems - Require concomitant therapy with any medications that pose safety issues - Medical history of medical disorders that would increase potential risk of study treatment or interfere with study participation - Had more than 7 days of inpatient treatment for substance use disorder in the 30 days prior to randomization - History of other psychoactive substance abuse or dependence (other than nicotine, cannabis, and habitual caffeine use) by DSM-IV criteria in the last 90 days (6 months for opiate abuse) or by urine drug screen - Abnormal AST or ALT (> 3 times of normal level) or elevated bilirubin - Pregnancy or nursing - Women of childbearing age not on an effective contraceptive method - Sensitivity to the study medications - Unstable medical conditions (e.g., serum liver enzyme levels > 3 times the upper limit of normal) - No contact person who could provide the whereabouts of the participant and without a fixed address or unavailable by phone or pager at the time of randomization - Illiteracy or unable to read English | - Concurrent DSM-IV criteria for depression, bipolar disorder, schizophrenia, bulimia/anorexia, dementia, anxiety disorders - Required psychotherapy - On antidepressants, mood stabilizers, antiepileptics or other medications - Use of any psychoactive drugs as evident by urine test in the last 30 days - Lifetime diagnosis of psychoactive substance dependence (except nicotine and coffee) - Medical conditions that would increase the potential risk of the study treatment or interfere with the study participation - Abnormal AST or ALT levels (>5 times of normal level) - Pregnancy or nursing - Women of childbearing age not on an effective contraceptive method - Sensitivity to the study medication - Illiteracy or unable to read German | - Any unstable active medical or additional psychiatric conditions - Active suicidal ideation - History of hypersensitivity or allergic reaction to acamprosate - Currently taking disulfiram - Currently being, or within the last 3 weeks having been treated with acamprosate - Abnormal ALT or AST levels (>3 times of normal level) - Diagnosis of primary biliary cirrhosis, chronic active hepatitis, and drug-induced hepatic insufficiency, as noted in the medical record - Moderate to severe renal impairment (creatinine level >1.5 mg/dL) - Pregnancy, plan of pregnancy in the next year, or nursing - Unable to provide informed consent - Unable to speak English |

**Supplementary Table S2:** Loci* with suggestive evidence of association (p<5E-06) with one of the outcome measures when analyzing the full cohort (all treatments)

| Outcome | rsID | chr:position | MA | CA | MAF | HR | Dir | P.value | Gene Annotation |
| --- | --- | --- | --- | --- | --- | --- | --- | --- | --- |
| Time Until Relapse | rs1078110 | 1:41252782 | C | G | 0.304 | 0.67 | --- | 6.2E-07 | intronic (KCNQ4[0]) |
|  | rs3097240 | 5:38605357 | G | A | 0.403 | 0.71 | --- | 9.9E-07 | ncRNA_intronic (LIFR-AS1[0]) |
|  | rs584789 | 5:118087436 | C | A | 0.303 | 1.37 | +++ | 2.0E-06 | intergenic (RP11-2N5.1[124]; RNU7-34P[6]) |
|  | rs77583603 | 2:186954132 | G | A | 0.089 | 1.69 | +++ | 2.0E-06 | intergenic (AC097500.2[6]; AC104058.1[74]) |
|  | rs433374 | 11:36994253 | C | A | 0.172 | 1.48 | +++ | 3.3E-06 | intergenic (CTD-2119L1.1[275]; SNORA31[729]) |
|  | rs79484822 | 4:70932193 | C | T | 0.095 | 1.60 | +++ | 4.5E-06 | upstream (CSN1S2AP[0]) |
|  | rs113018018 | 4:4887897 | A | G | 0.149 | 1.50 | +++ | 5.0E-06 | intergenic (MSX1[22]; LDHAP1[8]) |
| Time Until Heavy Relapse | **rs56951679** | **2:28483718** | **C** | **T** | **0.174** | **1.53** | **+++** | **1.6E-08** | **intronic (BRE[0])** |
|  | rs3019626 | 11:61808272 | t | c | 0.216 | 0.66 | --- | 1.7E-07 | intergenic (RP11-810P12.1[28]; RP11-810P12.6[32]) |
|  | rs2097759 | 17:36079453 | C | A | 0.153 | 0.63 | --- | 2.9E-06 | intronic (HNF1B[0]) |
|  | rs2015157 | 19:9346762 | G | T | 0.074 | 1.67 | +++ | 3.2E-06 | ncRNA_exonic (OR7D1P[0]) |
|  | rs11608708 | 12:132123936 | G | A | 0.188 | 1.46 | +++ | 3.5E-06 | intergenic (RP11-495K9.9[16]; RP11-495K9.7[8]) |

MA=minor allele; CA=common allele; MAF=minor allele frequency; HR=hazard ratio reported in terms of MA. HR>1 indicates a higher risk of relapse/heavy-relapse associated with an additional copy of the MA, whereas HR<1 indicates a lower risk. Dir=Direction of effect (HR>1 or HR<1) in the three contributing studies (ordered as: COMBINE/CITA/PREDICT)

*results were clumped using r^2^ of .1 within 250kb regions

**Supplementary Table S3:** Loci* with suggestive evidence of association (p<5E-06) with one of the outcome measures in analyses limited to patients treated with naltrexone

| Outcome | rsID | chr:position | MA | CA | MAF | HR | Dir | P.value | Gene Annotation |
| --- | --- | --- | --- | --- | --- | --- | --- | --- | --- |
| Time Until Relapse | rs62533259 | 9:9535958 | C | T | 0.140 | 2.20 | ++ | 7.7E-08 | intronic (PTPRD[0]) |
|  | rs11588477 | 1:228512036 | G | T | 0.223 | 1.87 | ++ | 8.6E-08 | intronic (OBSCN[0]) |
|  | rs7325923 | 13:24078596 | T | A | 0.109 | 2.14 | ++ | 2.1E-07 | intergenic (LINC00352[2]; TNFRSF19[66]) |
|  | rs1362196 | 7:34413651 | G | A | 0.058 | 2.75 | ++ | 9.3E-07 | ncRNA_intronic (NPSR1-AS1[0]) |
|  | rs117970302 | 15:88518599 | T | C | 0.094 | 2.13 | ++ | 1.2E-06 | intronic (NTRK3[0]) |
|  | rs11770131 | 7:23872506 | A | T | 0.070 | 2.42 | ++ | 1.3E-06 | downstream (STK31[0]) |
|  | rs114469564 | 21:43404351 | G | T | 0.050 | 2.57 | ++ | 1.7E-06 | intergenic (C2CD2[30]; ZBTB21[3]) |
|  | rs2908000 | 7:9898826 | C | T | 0.334 | 0.57 | -- | 1.8E-06 | intergenic (GS1-69O6.1[8]; AC006373.1[102]) |
|  | rs7828702 | 8:3003780 | G | A | 0.101 | 2.06 | ++ | 1.8E-06 | intronic (CSMD1[0]) |
|  | rs72765992 | 5:58579309 | C | G | 0.081 | 2.21 | ++ | 2.1E-06 | intronic (PDE4D[0]) |
|  | rs6076964 | 20:6210743 | T | C | 0.070 | 2.75 | ++ | 2.2E-06 | intergenic (AL109618.1[1]; CASC20[217]) |
|  | rs80081786 | 1:177797988 | C | T | 0.156 | 1.94 | ++ | 2.6E-06 | intergenic (RP11-63B19.1[119]; RP4-798P15.3[100]) |
|  | rs7141455 | 14:52227835 | T | C | 0.081 | 2.48 | ++ | 3.1E-06 | ncRNA_exonic (RP11-280K24.1[0]) |
|  | rs7187668 | 16:87077722 | T | C | 0.111 | 0.41 | -- | 3.5E-06 | intergenic (RP11-107C10.1[138]; RP11-134D3.1[14]) |
|  | rs12323856 | 14:105293185 | G | T | 0.051 | 6.54 | +? | 3.8E-06 | intergenic (LINC00638[3]; RPS26P49[4]) |
|  | rs11010438 | 10:36386254 | T | G | 0.123 | 1.84 | ++ | 4.8E-06 | intergenic (RP11-810B23.1[8]; MTND5P17[336]) |
| Time Until Heavy Relapse | **rs12749274** | **1:56883491** | **a** | **g** | **0.079** | **2.90** | **++** | **3.9E-08** | **intergenic (RP4-710M16.2[2]; PPAP2B[77])** |
|  | rs62533259 | 9:9535958 | C | T | 0.138 | 2.36 | ++ | 6.1E-08 | intronic (PTPRD[0]) |
|  | rs7141455 | 14:52227835 | t | c | 0.081 | 2.70 | ++ | 4.8E-07 | ncRNA_exonic (RP11-280K24.1[0]) |
|  | rs71569385 | 6:37575654 | T | C | 0.050 | 3.14 | ++ | 7.6E-07 | intergenic (MIR4462[52]; MDGA1[25]) |
|  | rs78950771 | 5:154582312 | G | A | 0.070 | 2.73 | ++ | 1.9E-06 | intergenic (CTD-2311A18.1[90]; CTC-447K7.1[195]) |
|  | rs148035692 | 16:5143629 | a | g | 0.069 | 4.57 | ?+ | 2.0E-06 | intronic (FAM86A[0]) |
|  | rs57326784 | 1:209836879 | T | C | 0.065 | 3.70 | -+ | 2.0E-06 | ncRNA_intronic (RP1-28O10.1[0]) |
|  | rs11588477 | 1:228512036 | G | T | 0.225 | 1.79 | ++ | 2.0E-06 | intronic (OBSCN[0]) |
|  | rs34428249 | 10:88763843 | G | A | 0.075 | 2.46 | ++ | 2.8E-06 | ncRNA_intronic (AGAP11[0]; RP11-96C23.11[0];  RP11-96C23.5[0]; RP11-96C23.14[0]) |
|  | rs10788250 | 10:123901712 | t | c | 0.080 | 2.30 | ++ | 4.7E-06 | intronic (TACC2[0]) |

MA=minor allele; CA=common allele; MAF=minor allele frequency; HR=hazard ratio reported in terms of MA. HR>1 indicates a higher risk of relapse/heavy-relapse associated with an additional copy of the MA, whereas HR<1 indicates a lower risk. Dir=Direction of effect (HR>1 or HR<1) in the three contributing studies (ordered as: COMBINE/PREDICT)

*results were clumped using r^2^ of .1 within 250kb regions

**Supplementary Table S4:** Loci* with suggestive evidence of association (p<5E-06) with one of the outcome measures in analyses limited to patients treated with acamprosate

| Outcome | rsID | chr:position | MA | CA | MAF | HR | Dir | P.value | Gene Annotation |
| --- | --- | --- | --- | --- | --- | --- | --- | --- | --- |
| Time Until Relapse | **rs77583603** | **2:186954132** | **G** | **A** | **0.093** | **2.38** | **+++** | **3.1E-09** | **intergenic (AC097500.2[6]; AC104058.1[74])** |
|  | rs78024421 | 13:58883475 | C | T | 0.059 | 2.52 | +++ | 8.8E-07 | intergenic (LINC00374[76]; RNY4P29[218]) |
|  | rs4582229 | 5:60848745 | T | C | 0.211 | 0.56 | --- | 1.1E-06 | intergenic (ZSWIM6[7]; AC008836.1[4]) |
|  | rs7233031 | 18:5480461 | G | A | 0.271 | 1.57 | +++ | 1.9E-06 | ncRNA_intronic (RP11-286N3.1[0]) |
|  | rs2435356 | 10:43583150 | A | G | 0.250 | 1.56 | +++ | 2.9E-06 | intronic (RET[0]) |
|  | rs10965428 | 9:22718481 | C | A | 0.086 | 1.90 | +++ | 3.0E-06 | ncRNA_intronic (RP11-399D6.2[0]) |
|  | rs11601116 | 11:36986344 | T | C | 0.187 | 1.63 | +++ | 3.0E-06 | intergenic (CTD-2119L1.1[267]; SNORA31[737]) |
|  | rs72684759 | 14:21889563 | T | C | 0.129 | 2.14 | +?+ | 3.2E-06 | intronic (CHD8[0]) |
|  | rs27565 | 5:59837591 | C | T | 0.468 | 0.67 | --- | 3.5E-06 | ncRNA_intronic (PART1[0]) |
|  | rs7249601 | 19:57546158 | C | T | 0.066 | 2.03 | +++ | 4.0E-06 | intergenic (CTC-258N23.3[113]; RPL7AP69[31]) |
|  | rs213041 | 1:21657769 | T | C | 0.087 | 1.87 | +++ | 4.2E-06 | intronic (ECE1[0]) |
|  | rs702131 | 9:12526192 | G | A | 0.279 | 1.54 | +++ | 4.2E-06 | intergenic (RNU2-47P[226]; TYRP1[159]) |
|  | rs35734605 | 13:50149649 | T | C | 0.094 | 2.43 | +?+ | 4.5E-06 | intronic (RCBTB1[0]) |
|  | rs9924363 | 16:19686316 | C | A | 0.485 | 0.66 | --- | 4.9E-06 | intronic (C16orf62[0]) |
| Time Until Heavy Relapse | rs34797278 | 17:14311624 | G | A | 0.079 | 2.14 | +++ | 5.4E-08 | ncRNA_intronic (AC022816.2[0]) |
|  | rs79234780 | 10:82585091 | A | G | 0.052 | 2.24 | +++ | 3.5E-07 | intergenic (FARSBP1[47]; WARS2P1[115]) |
|  | rs113943471 | 6:88519981 | t | c | 0.077 | 2.85 | +?+ | 4.8E-07 | intergenic (snoU13[89]; Y_RNA[16]) |
|  | rs6087091 | 20:1050896 | t | c | 0.234 | 1.57 | +++ | 1.4E-06 | intergenic (RSPO4[68]; PSMF1[43]) |
|  | rs12702045 | 7:43507689 | a | c | 0.222 | 1.66 | +++ | 1.4E-06 | intronic (HECW1[0]) |
|  | rs10843660 | 12:30368457 | t | c | 0.421 | 0.65 | --- | 1.6E-06 | ncRNA_intronic (RP11-776A13.1[0]) |
|  | rs6865889 | 5:123683057 | t | c | 0.084 | 1.92 | +++ | 1.6E-06 | ncRNA_intronic (LINC01170[0]) |
|  | rs17160835 | 7:138948598 | G | A | 0.197 | 1.59 | +++ | 2.1E-06 | intronic (UBN2[0]) |
|  | rs400781 | 19:53138219 | A | G | 0.055 | 2.24 | +++ | 2.2E-06 | intronic (ZNF83[0]) |
|  | rs13298120 | 9:36640907 | t | c | 0.165 | 1.68 | +++ | 2.6E-06 | intronic (MELK[0]) |
|  | rs288485 | 7:130450214 | G | A | 0.052 | 2.22 | +++ | 2.7E-06 | intergenic (KLF14[31]; RP11-36B6.1[25]) |
|  | rs75466853 | 13:59395212 | G | A | 0.065 | 2.09 | +++ | 3.1E-06 | intergenic (DNAJA1P1[69]; HMGN2P39[189]) |
|  | rs73650194 | 9:22749908 | T | A | 0.057 | 2.04 | +++ | 3.3E-06 | ncRNA_intronic (RP11-399D6.2[0]) |
|  | rs56010289 | 7:28468621 | G | C | 0.068 | 2.14 | +++ | 3.6E-06 | intronic (CREB5[0]) |
|  | rs2435356 | 10:43583150 | a | g | 0.246 | 1.55 | +++ | 3.9E-06 | intronic (RET[0]) |
|  | rs1335855 | 1:34648046 | C | T | 0.129 | 1.68 | +++ | 4.5E-06 | intronic (C1orf94[0]) |
| MA=minor allele; CA=common allele; MAF=minor allele frequency; HR=hazard ratio reported in terms of MA. HR>1 indicates a higher risk of relapse/heavy-relapse associated with an additional copy of the MA, whereas HR<1 indicates a lower risk. Dir=Direction of effect (HR>1 or HR<1) in the three contributing studies (ordered as: COMBINE/CITA/PREDICT). *results have been clumped using r^2^ of .1 within 250kb regions | | | | | | | | | |

**Supplemental Figure S1:** QQ plots for meta-analyses of (A) Time until Relapse and (B) Time until Heavy Relapse, in the full cohorts (all treatments). Manhattan plots for the corresponding analyses are shown in Figure 1.

B

A

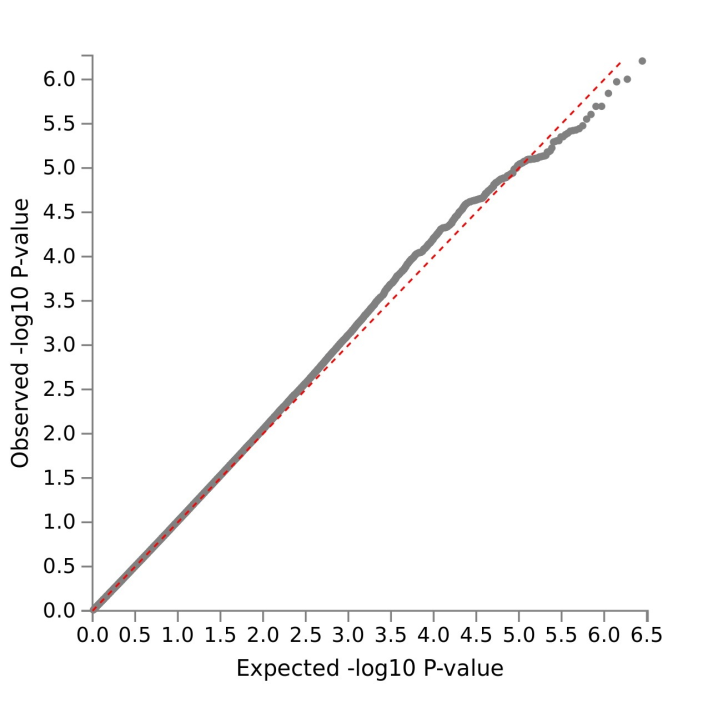

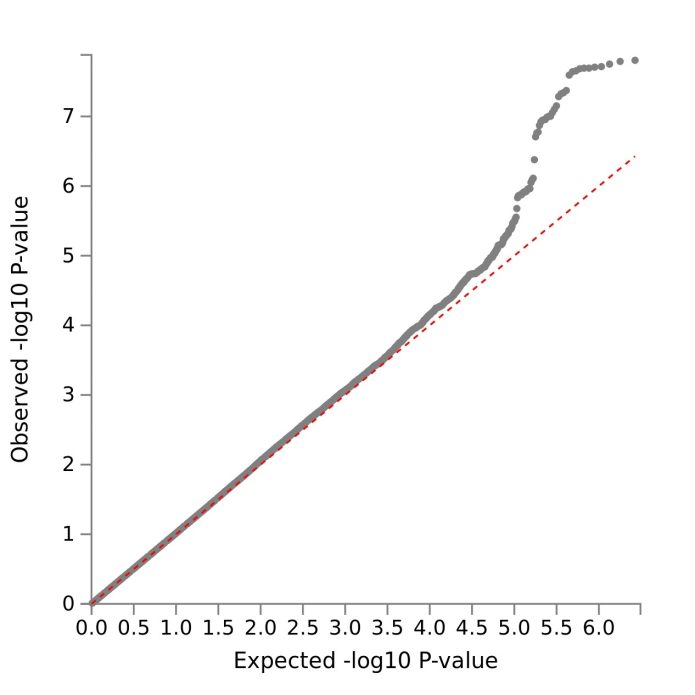

**Supplemental Figure S2**: Regional association plot for the BRE locus in the full cohort analysis of time until heavy relapse, generated using LocusZoom [4].

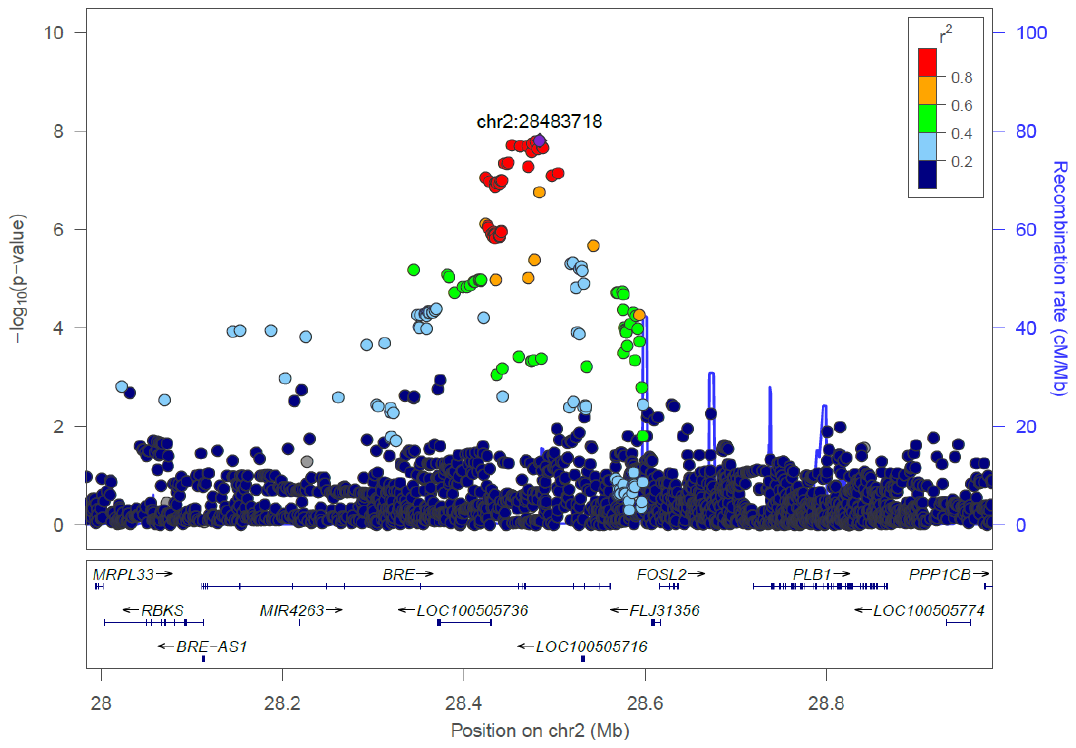

**Supplemental Figure S3:** Manhattan Plots for (A) TR and (B) THR in naltrexone-treated subjects. The corresponding QQ plots for TR and THR are shown in panels (C) and (D), respectively.

A

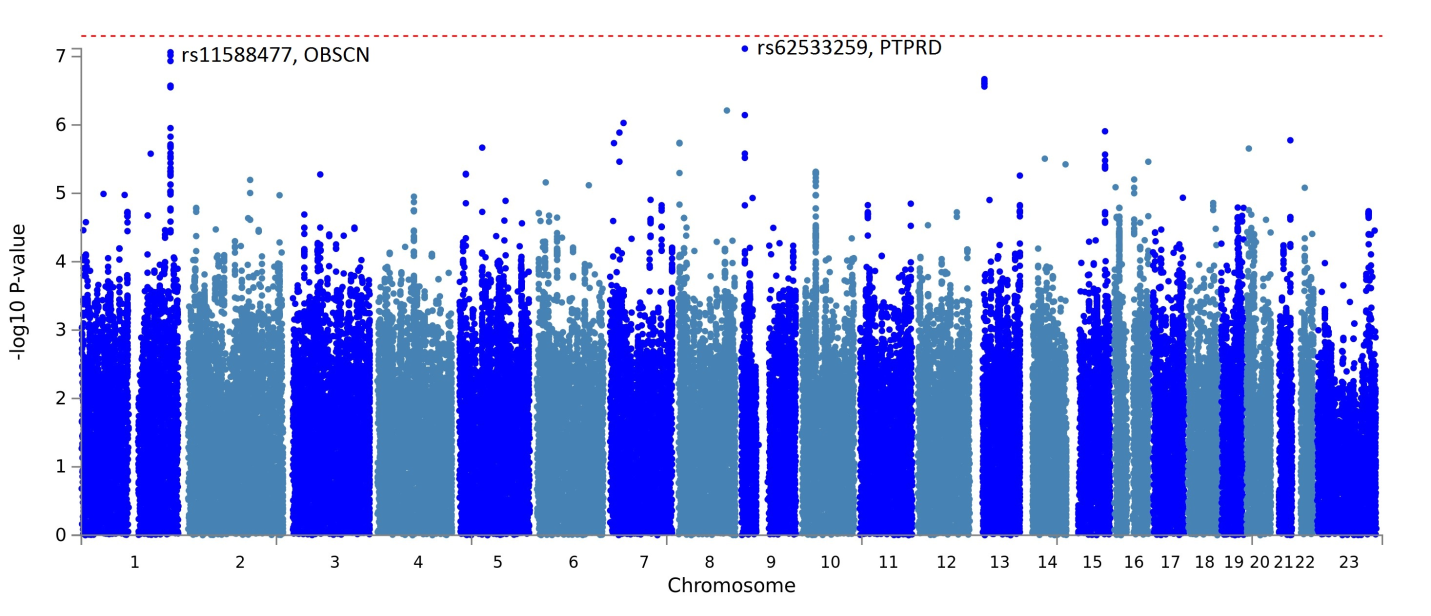

B

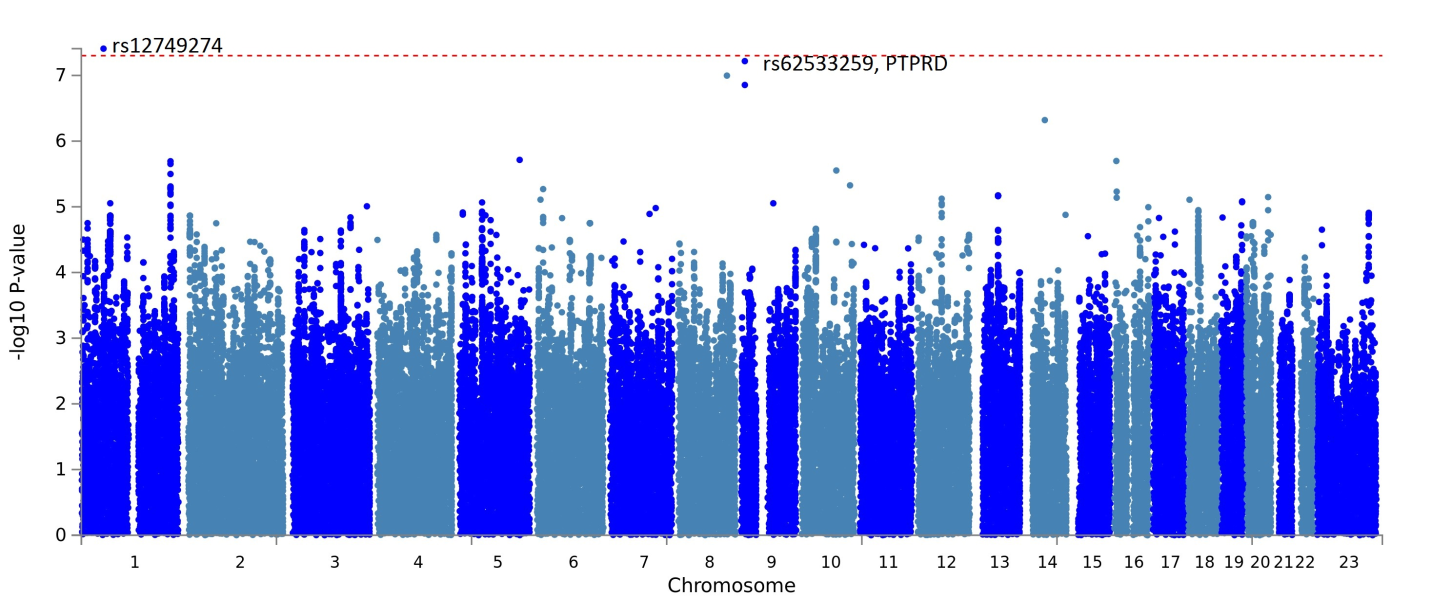

C	D

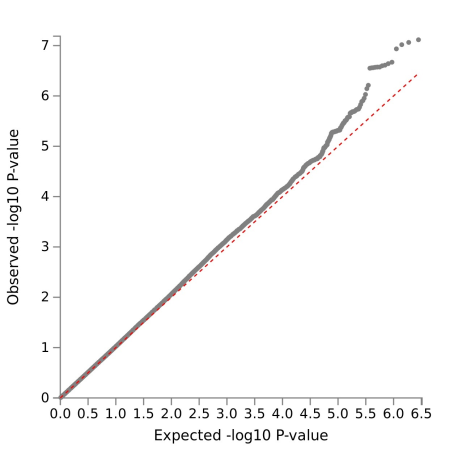

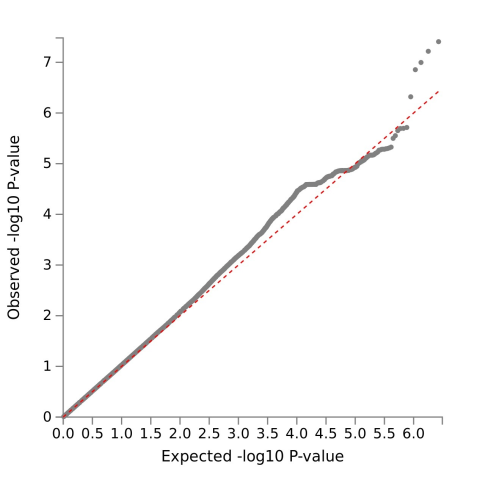

**Supplemental Figure S4:** Manhattan Plots for (A) TR and (B) THR in acamprosate-treated subjects. The corresponding QQ plots for TR and THR are shown in panels (C) and (D), respectively.

A

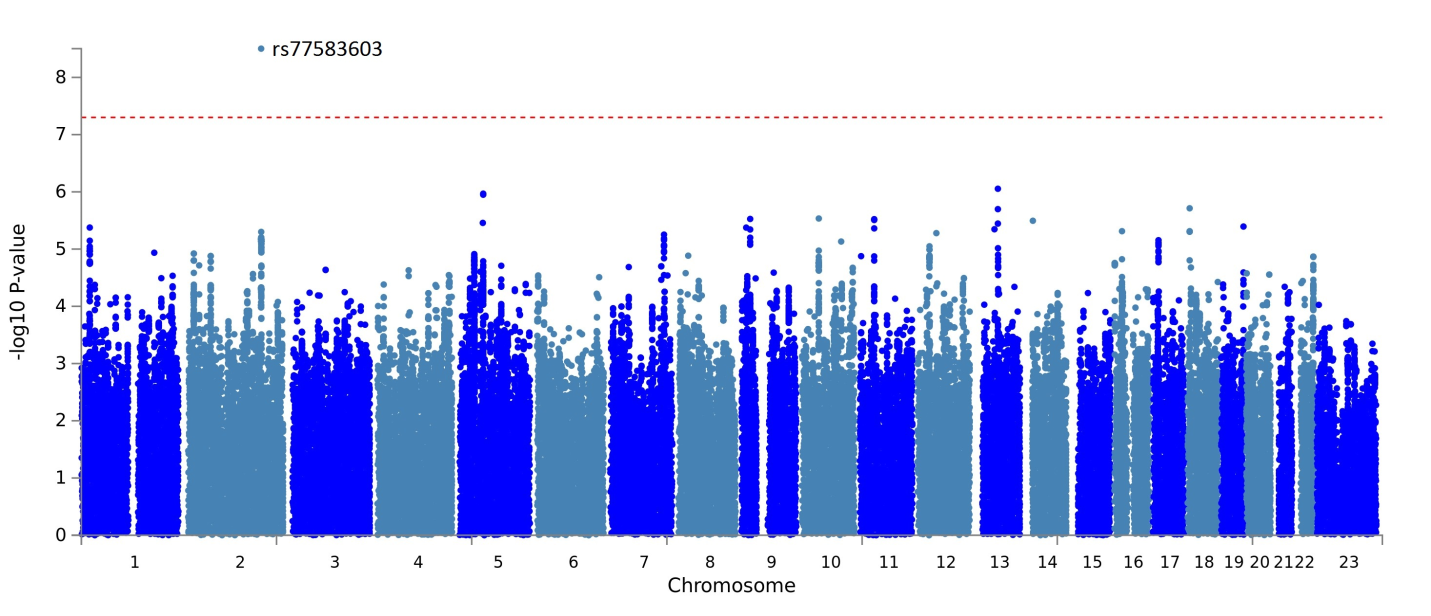

B
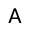

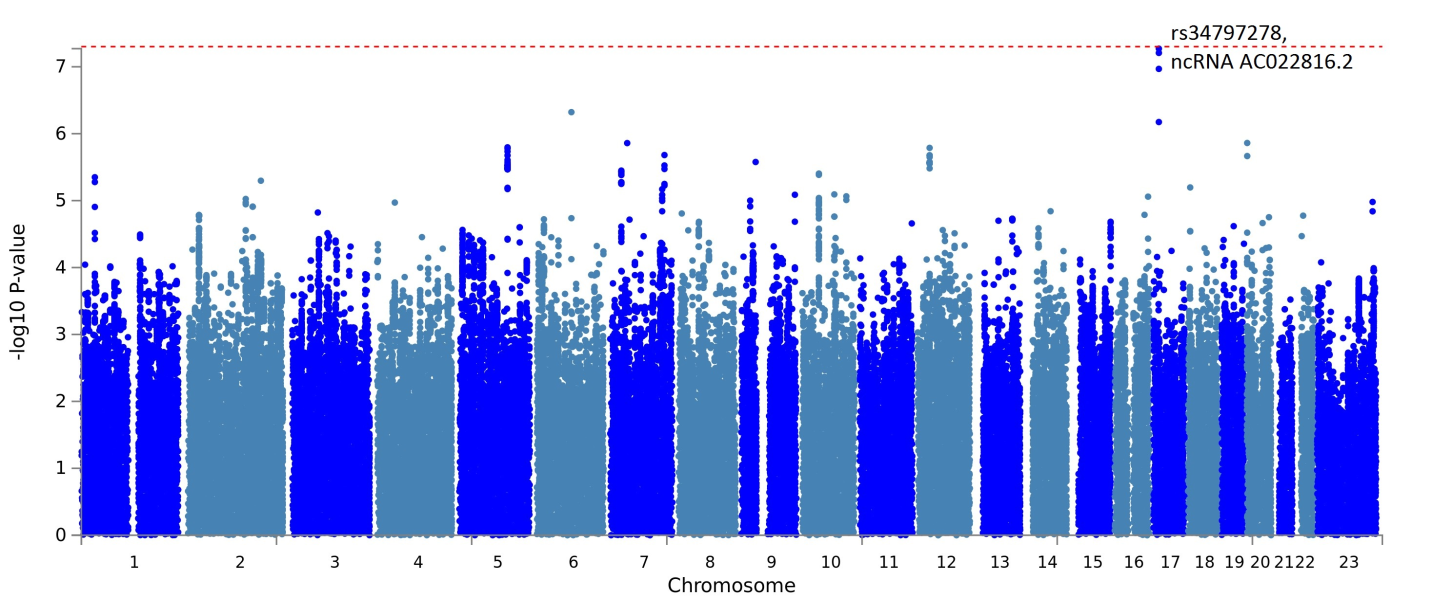

C	D

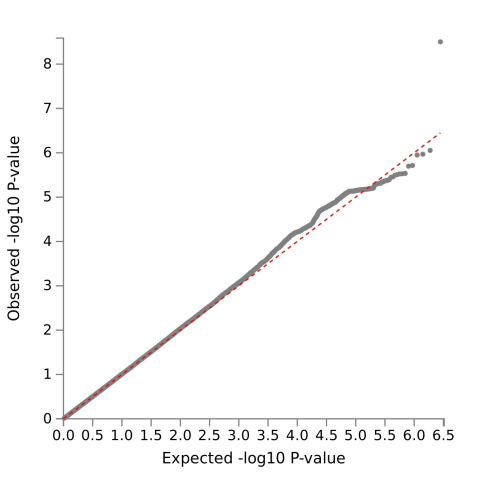

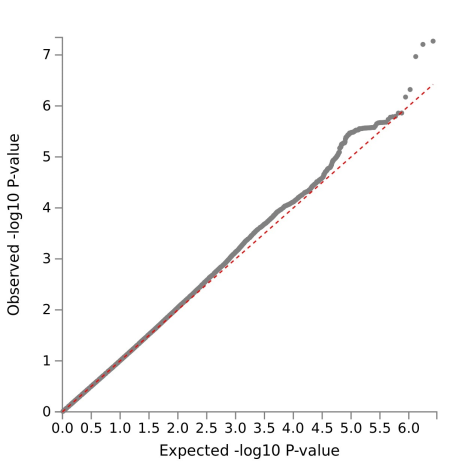

**Supplemental Figure S5**: Scatterplots comparing results (p-values) from GWAS of TR vs. THR, and comparing results of analyses of different patient subsets. Scatterplots of –log(p) for (A) analysis of TR vs. analysis of THR in all subjects, (B) analysis of TR vs. analysis of THR in naltrexone-treated subjects, (C) analysis of TR vs. analysis of THR in acamprosate-treated subjects, (D) analysis of TR in acamprosate-treated patients vs. analysis of TR in naltrexone-treated patients, and (E) analysis of THR in acamprosate-treated patients vs. analysis of THR in naltrexone-treated patients

A	B	C

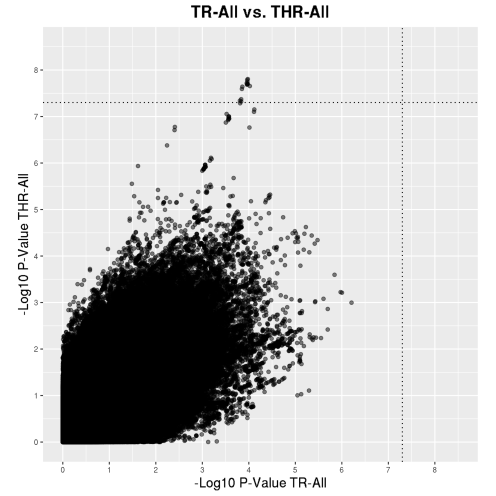

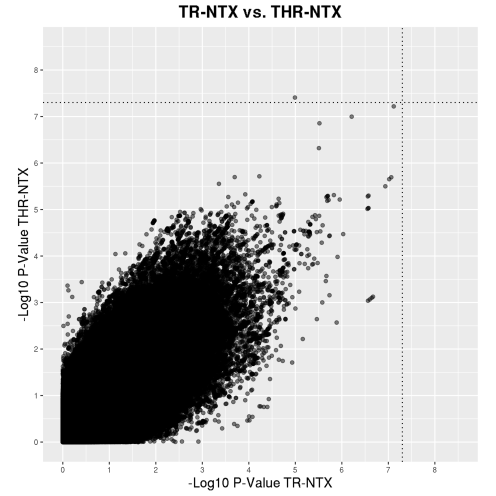

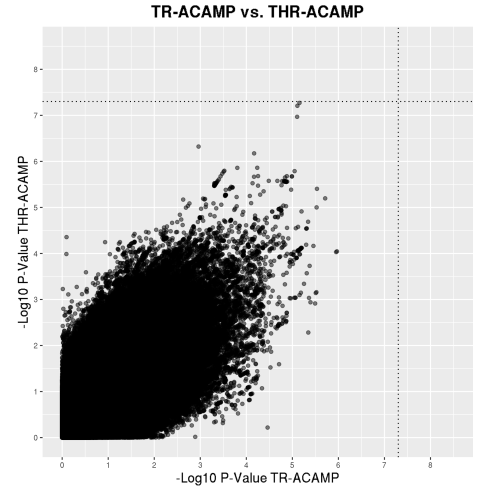

PTPRD rs62533259

BRE rs56951679

**D	E**

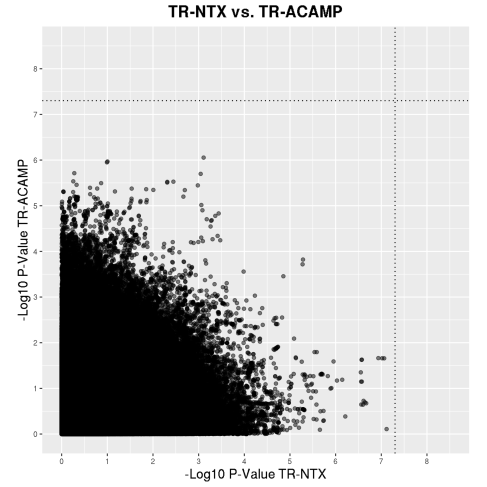

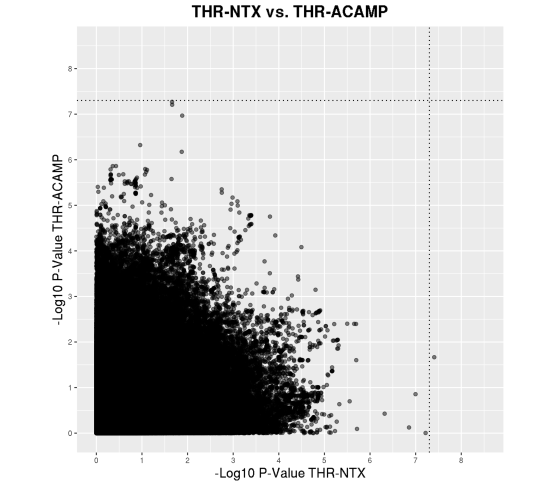

PTPRD rs62533259

PTPRD rs62533259

**Supplemental Figure S6:** MAGMA tissue enrichment analysis in the naltrexone-treated patients. This analysis tests for enrichment of differentially expressed gene sets in a given tissue compared to all other tissue types. The -log10(p-value) for the enrichment analysis of each analyzed tissue is shown on the y-axis.

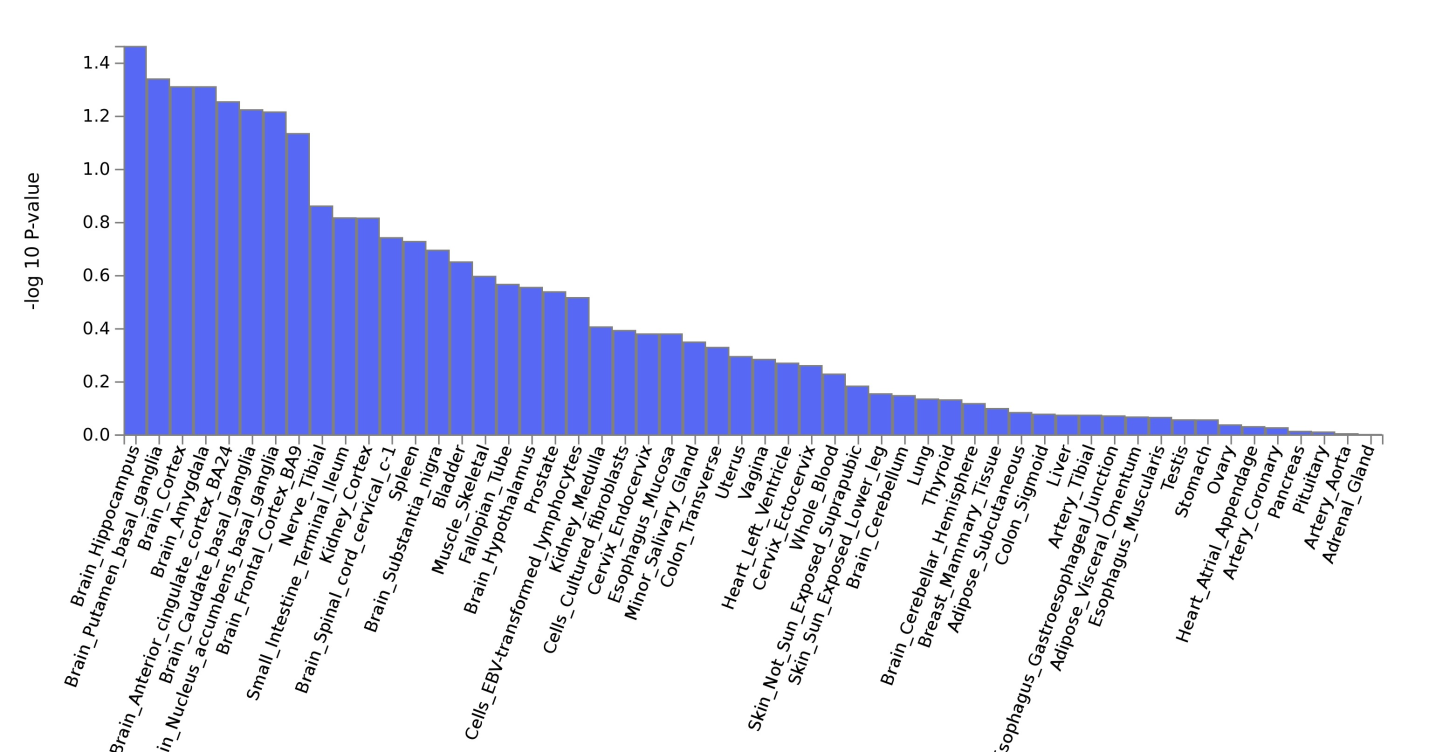

References

1.	Treutlein, J., et al., *Genome-wide association study of alcohol dependence.* Arch Gen Psychiatry, 2009. **66**(7): p. 773-84.

2.	Frank, J., et al., *Genome-wide significant association between alcohol dependence and a variant in the ADH gene cluster.* Addict Biol, 2012. **17**(1): p. 171-80.

3.	Pritchard, J.K., M. Stephens, and P. Donnelly, *Inference of population structure using multilocus genotype data.* Genetics, 2000. **155**(2): p. 945-59.

4.	Pruim, R.J., et al., *LocusZoom: regional visualization of genome-wide association scan results.* Bioinformatics, 2010. **26**(18): p. 2336-7.
